# Using high *effective risk* of Adult-Senior duo in multigenerational homes to prioritize COVID-19 vaccination

**DOI:** 10.1101/2021.04.14.21255468

**Authors:** Brijesh Saraswat, Santosh Ansumali, Meher K. Prakash

## Abstract

Universal vaccination on an urgent basis is a way of controlling the COVID-19 infections and deaths. Shortages of vaccine supplies and practical deployment rates on the field necessitate prioritization. The *global* strategy has been to prioritize those with a high personal risk due to their age or comorbidities and those who constitute the essential workforce of the society. Rather than a systematic age-based roll-down, assigning the next priority requires a *local* strategy based on the vaccine availability, the effectiveness of these specific vaccines, the population size as well as its age-demographics, the scenario of how the pandemic is likely to develop. The Adult (ages 20-60) – Senior (ages over 60) duo from a multigenerational home presents a high-risk demographic, with an estimated “effective age” of an adult living with a grandparent that is not vaccinated to be 40 years more. Our model suggests that strategically vaccinating the Adults from multigenerational homes in India may be effective in saving the lives of around 70,000 to 200,000 of Seniors, under the different epidemiological scenarios possible with or without strict lockdowns.

## Introduction

COVID-19 pandemic management required the use of Non-Pharmaceutical Interventions (NPIs) including severe full or graded lockdowns [Ferguson *et al*. 2020]. The nearly 3 million deaths caused by COVID-19 across the world are much lesser than the counterfactual scenario with no interventions. Although herd-immunity through infection was seen as an option, the unprecedented scale of deaths argued against it [Randolph *et al*. 2020; Fontanet *et al*. 2020]. Thus the NPIs continue to play a critical role of flattening the infection curve [Anderson *et al*. 2020].

The only way of ending the pandemic, and to end the cycle of graded lockdowns is immediate and universal vaccination. Commensurate with the scale of the pandemic, several vaccines have been developed and became available at an accelerated speed [Corum *et al*. 2020; Forni *et al*. 2021; Voysey *et al*. 2020]. These vaccines have demonstrated effectiveness ranging from 60% to 95% [Forni *et al*. 2021]. Although these vaccines are available and effective in principle, there are several challenges in their deployment. The vaccines based on mRNA technology require a cold-chain of −80°C, while others with less stringent cold-chain requirements have lower effectiveness. More importantly, all of these vaccines are with a limited production and supply capacity [Mills and Salisbury, 2020; Irwin 2021] compared to the immediate global need.

Considering these challenges, the taskforces across the world have recommended prioritized vaccination of the vulnerable [NASEM, 2020]. The principal factor guiding the recommendations is the mortality risk posed by the age (over 60 years) and comorbidities. The priority group also included health care workers and other essential work force of the society, who although may not be at the risk of death, cannot be quarantined due to infections without crippling the functional society. The choice of the vulnerable and the essential for the first priority was almost universally adopted by all the governments.

However, the strategy for the next phase of prioritization is not obvious. At the rate of current vaccination, it is feared that most countries, may need at least another year in the ideal scenario without any manufacturing or distribution delays. A systematic roll-down across the different age groups is not a practical strategy for many countries, especially if they have a large young population, such as in India. The ‘pyramid’ structure of the age demographics in the population, where the population increases in the younger age brackets, presents more candidates to be vaccinated in each roll-down phase. Paradoxically, the younger population which is larger in number also has a lower mortality risk [Levin et al. 2020; Ghisolfi *et al*. 2020], which requires a re-examination of the vaccination strategy beyond those covered in the first priority.

Since the availability of vaccines in USA has not been a challenge, and most European countries are still vaccinating those in the first priority category, it appears that the grey area that is faced by India today on “Who is next?”, did not receive sufficient attention. This may be a question that the European countries may be confronted with as soon as the first priority group is vaccinated.

Vaccinating the family members of the vulnerable is a well-known strategy when the vaccine is considered not so safe or if the vulnerable have a lower immunity levels, due to a pre-existing HIV infection or other factors which prevent them from developing antibodies [Public Health England, 29 March 2021]. However, since the safety of all the COVID-19 vaccines has been demonstrated, the discussion of vaccinating the family members of the vulnerable did not take prime importance.

In this work, we systematically evaluate and demonstrate the effectiveness of this strategy of vaccinating the young family members from the multigenerational homes on a priority basis in the Indian context, by combining it with the local realities of vaccination availability, effectiveness as well as the developing epidemiological scenario.

## Results and Discussion

### Indian COVID-19 and vaccination scenario

The vaccination strategy should consider the three *local* realities of the society for which the analysis is meant: Vaccine availability, COVID-19 incidence, age-demographics.

#### Vaccine availability

COVISHIELD (Serum Institute of India-AstraZeneca Plc) [Voysey *et al*. 2020] and COVAXIN (Bharat Biotech Intl. Ltd.) [Ella *et al*. 2021] are the first two vaccines that were approved for use in India. Both of these vaccines were proven for their safety. The former has shown an effectiveness of 67% in reducing infections, 70% in reducing transmission [Voysey *et al*. 2021], and we assume the same for the latter. We further assume an 80% reduction in age-dependent mortality due to the vaccination [Bernal *et al*. 2021; Public Health England 01 March 2021]. The combined production capacity of these two vaccines is around 60 million doses monthly. A step-up in production is expected between July to September 2021, but we present our model based on the foreseeable situation in mid-April.

#### COVID-19 incidence

Flu-like infections spread rapidly in winter. In the winter of 2020, India was in an ‘unlock’ phase with minimal NPIs, and yet COVID-19 infections were contained. Several hypotheses surrounded this observation - including protection from previous infectious diseases, herd immunity with a large unmonitored COVID-19 infection. However, in the spring of 2021, the second wave of COVID-19 started, with case-loads exceeding 150,000 daily infections (*https://www.covid19india.org*). The rising pattern appears to parallel the rise of infections in the spring and summer of 2020. If this situation continues, either because there is an inherent spread of infection under hot conditions due to climatization [Sruthi *et al*. 2021] or due to any other reasons, one may expect rising infections until September 2021.

Typically one uses Susceptible-Infected-Recovered (SIR) [Kermack and McKendrick, 1927] or Susceptible-Asymptomatic-Infected-Recovered (SAIR) [Kaushal *et al*. 2020] models to make predictions of COVID-19 incidence. In fact, much simpler extrapolations based on global trends were also successful in making reliable predictions for a few weeks [Prakash *et al*. 2020]. However, in order to forecast for many months, one has to assume various conditions that allow COVID-19 to spread - for example, the transmission rate or the R_0_, how it will be managed by lockdowns. Since making an assumption is unavoidable, our assumptions for the second wave were guided by the global trends in the peak of infections and duration of the second-wave. The motivation for this assumption was that unless the susceptible population is completely exhausted, the peak is ultimately defined by the lockdowns as a response to a breakdown of the healthcare infrastructure.

Studying the second-wave from the countries [Worldometers, 2020] which had the most infections (**Supplementary Table 1**), it is clear that all of them had a peak of the second wave that was worse than the first wave of the spring of 2020 (ratio: 4.35±1.95). Several factors may have contributed to this large second peak in most countries, including the need to keep their economies active as well as their confidence in a better management of critical cases through effective treatment. Further, the *full-width at half-maximum (*a model-free way of defining the width) for the duration of the second wave in most COVID-19 affected countries lasted 27 to 122 days, with extreme and moderate restrictive NPIs respectively. This duration is also commensurate with the width of half-maximum (108 days) of the first wave of COVID-19 infections which occurred in the same season in 2020. While we acknowledge that it is hard to predict whether the actual trends in infections which may be checked by several unknowns such as attainment of herd-immunity, or imposition different grades of lockdowns, the global trends in the peak and the width of the second-wave serve as a guideline for estimating the consequences under some realistic worst-case scenarios (**Population Level Model** below).

#### Age demographics

The age-binned population data was obtained from the 2011 census. To estimate the population distribution in 2021, the population in each bin was multiplied by 1.125 to match the overall 12.5% increase in the Indian population between 2011-2021. It is evident that the population is largely young: 553 million (ages 0-20), 527 million (20-45), 173 million (45-60), 119 million (60+), with a low individual risk and requires multiple layers of prioritization.

### Population level Model

#### Focus

The focus of the work is on how vaccination of the “Adults” (ages 20 to 60) reduces the secondary infections and deaths among the “Seniors” (ages 60 and above). The deaths among the Adults, and the deaths among Seniors who were infected by means other than through families are *not* analyzed or reported.

#### Contact structure

The home-contact matrix for India [Prem *et al*. 2017] shows a banded structure with a contact between same age group, and those with differences of 25 and 50 years, suggesting an underlying generational gap of 25 years. We assume each Senior of age *S* is related to one child of age (*S-25*) and one grandchild of age (*S-50*). Since the focus is on intergenerational infections, this assignment is performed only if the age of the child or the grandchild is <60.

#### Overall incidence

We consider four different scenarios of the development of COVID-19 second wave in India. The number of daily infections crossed 150,000 by 12 April 2021. We considered four different scenarios starting from this. Scenario 1, was meant to be a conservative scenario of infections reaching a peak of 225,000 with a flattening for a month and then a subsequent decrease. In Scenarios 2 to 4, we considered a peak daily infection of 400,000, commensurate with the average ratio of the peak in the second versus the first waves in the top COVID-19 affected countries. Similarly, guided by the global average of the full width at half-peak with and without lockdowns (**Supplementary Table 1**), we assumed widths of around 60, 90 and 120 days for the Scenarios 2, 3, and 4 respectively of the second wave in India. Based on these assumptions, the four different epidemiological scenarios shown graphically in **Figure 2A**. While assumptions of the epidemiological scenarios in a model-independent way may seem *ad hoc*, qualitatively these are not very different from the *ad hoc* assumptions of the sequence of increase or decrease in R_0_ in the SIR/SAIR models.

**Figure 1.**
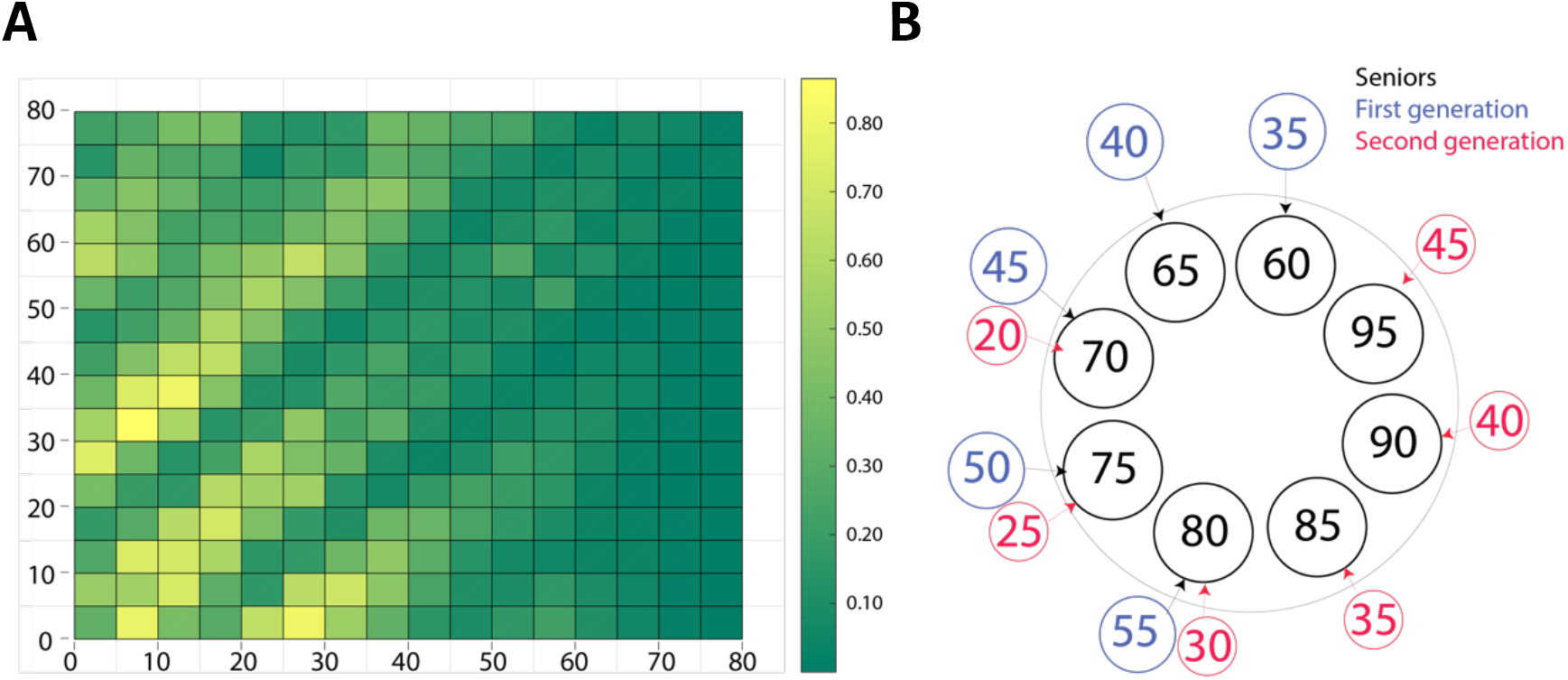
Intergenerational contact structure. **A**. Age-structured home contact network in India, obtained from Prem *et al*. 2017. The figure shows a clear banded structure, with a 25 year intergenerational time. The colorbar indicates the estimated mean number of contacts made by an individual (Prem *et al*. 2017) **B**. The inferred intra home contact network that was used in this work. The model assigns to every Senior of age *S>60*, a first generation home contact of *S-25* and a second generation home contact of *S-50*, as long as these contacts have an age of less than 60.

**Figure 2.**
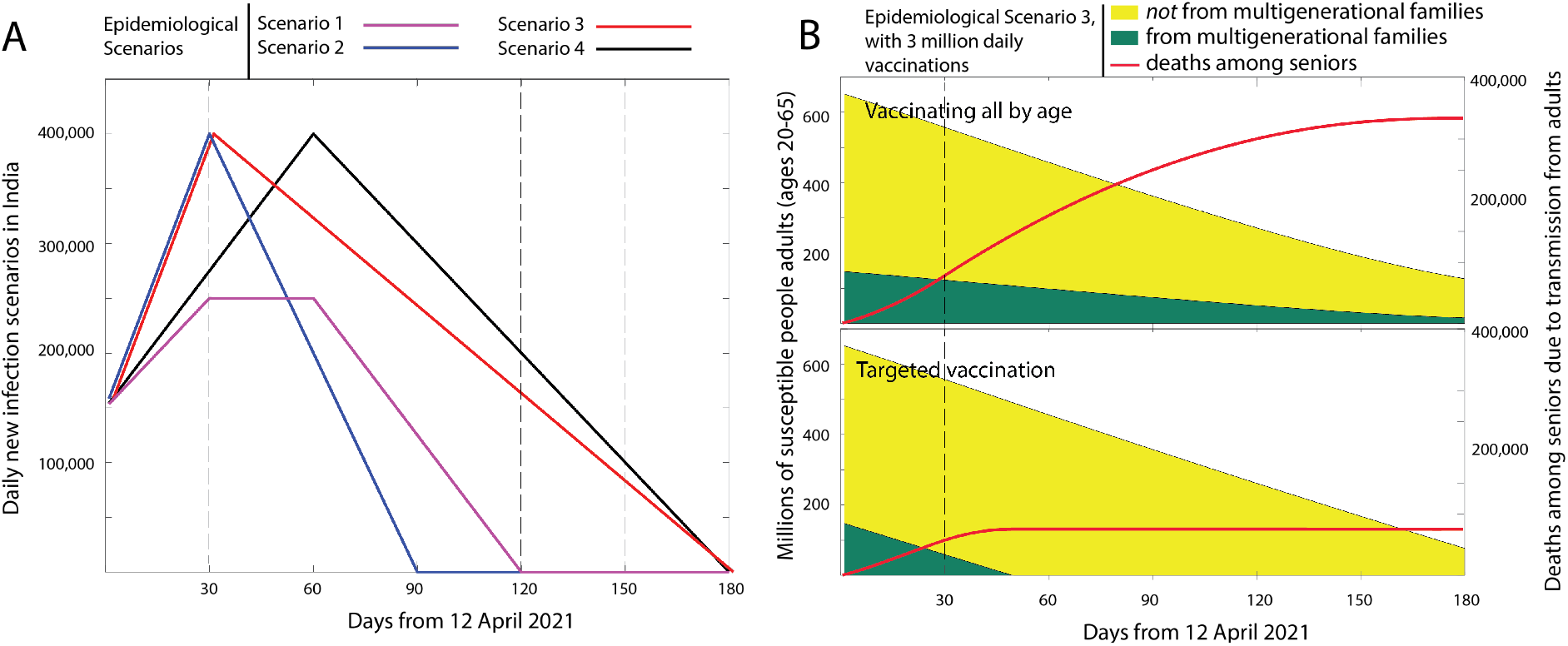
COVID-19 development scenarios and deaths avoided. **A**. The four COVID-19 development scenarios considered in this study. Scenario 1 is the most conservative scenario considering the daily infections on 12 April 2021. The peak of daily infections in all other scenarios is assumed to be 400,000 and is based on the average ratio of second to first peak observed in countries with the most infections (**Supplementary Table 1**). **B**. Comparison of two strategies based on (top) no-selectivity in vaccinating members of multigenerational families and (bottom) preferential vaccination to younger members of multigenerational families for scenario 3. The deaths reduced among the Seniors by vaccinating the adults in all four scenarios is shown in **Table 1**.

The total number of daily infections are assumed to be distributed in the population in the ratio of the susceptible population in each bin. The overall incidence so assumed serves to define the baseline infection rate for the unvaccinated Adults. However, the infections in the model differ from this. Among the Adults, the baseline number is corrected by the vaccinated fraction and the chances of infection despite the vaccination. Similarly, the infections transmitted by the Adults to the Seniors also change due to either or both of them being vaccinated.

#### Secondary attack rate

The average secondary attack due to a primary infection in the family is around 30% [Madewell *et al*., 2020].

#### Mortality

Since we examine the age-based vaccine rollout strategy, COVID-19 mortality is attributed to the Individual risk posed by the age [Levin et al. 2020; Ghisolfi et al. 2020]. People with comorbidities that increase the risk of COVID-19 fatality are considered to be vaccinated early on and independent of the age based roll out strategy. In the absence of the relevant data, it is assumed that the age dependent mortality does not change with the variants.

#### Effect of vaccination

The effect of vaccination is assumed mainly based on the data from AstraZeneca reports – the chance of infection is reduced by 67%, transmission by 70%, and mortality among the infected by 80%. We assume that these reductions are universally applicable to all age groups, and regardless of the vaccine type.

#### Other model assumptions

The many existing and possible new variants are considered not to be different, with respect to the fatality rates or for the effectiveness of the vaccine. At least in the 6 month period, a person who is infected with COVID-19, is assumed not to be reinfected. The potential deaths in Seniors are assigned to the day on which an adult is infected for simplicity of the analysis, although the same analysis may equally be presented with a time delay including hospitalization times. The vaccination rates may increase slightly, this increase may be compensated by the need for the second dose. Thus, to not invoke complicated scenarios of vaccine availability, hesitancy, second dose delays, we perform the analysis believing the 1^st^ dose of the vaccine implicitly delivers its effectiveness.

#### Calculations

Briefly, the calculations involve three different considerations, as shown in the **Supplementary Scheme 1** –

1. Initialization - the demographics of the population and assignment of some of the Adults to Multigenerational homes were performed while maintaining the intergenerational gap of 25 years. The vaccination status of the Adults and Seniors is defined based on the reported coverage [*https://dashboard.cowin.gov.in/]*.
2. Infection and Vaccination – the evolution of the COVID-19 case load, feasible vaccination rates and the choice of a targeted strategy and vaccination status of the Adults is updated on a daily basis in the model. The vaccinated as well as the infected are removed from the susceptible population.
3. Consequences to Seniors – only the infections transmitted from Adults to Seniors in multigenerational families are considered. The transmission, contraction and death rates are all influenced by the vaccination.

The calculations were performed on this population over a period of 180 days. The complete analysis code is available online (https://github.com/meherpr/vaccinating-young).

### Impact of prioritizing adults in a multigenerational home

By 12 April 2021, as shown in **Supplementary Table 3**, the priority vaccination for Seniors had a coverage of around 50%, and about 20% from the second priority group of ages 45-60 have been vaccinated [*https://dashboard.cowin.gov.in/]*. Under the four scenarios of COVID-19 second wave development (**Figure 2A**), we studied the effectiveness of vaccinating with two different strategies – age based roll-down to younger populations, and a preferential targeted vaccination of younger members from multigenerational homes. By adopting a targeted strategy, the susceptible Adults from multigenerational families may all be vaccinated in a relatively short time of 52 days if administered at 2 million doses per day, compared to a uniform age-based strategy (**Figure 2B**). In a more realistic scenario, the number of deaths among the Seniors due to transmissions from the adults in the family (**Table 1**) are likely to be reduced by over 100,000 due to this targeted strategy.

### Interpreting through Effective Risk and Effective Age

While the quantitative effectiveness of vaccinating Adults from multigenerational homes differs in the various epidemiological scenarios, the advantage remains in all scenarios and may be interpreted in a simpler way, by defining the “effective risk”. Age and the presence of comorbidities are individual risk factors for COVID-19 fatality [Levin et al. 2020; Ghisolfi et al. 2020]. The risk profile of an individual is defined by the infection fatality rate (IFR), which quantifies the chance of death when a person of a given demographic is infected by COVID-19. Defining an adult living in a multigenerational home as a new demographic allows us to obtain an “effective” risk profile for this group. The chance of death in this Adult-Senior duo, due to a COVID-19 infection in the Adult, is the chance of fatality in the Adult, combined with the chance of fatality in the Senior weighted by the chance of Secondary Attack (transmission) at home:

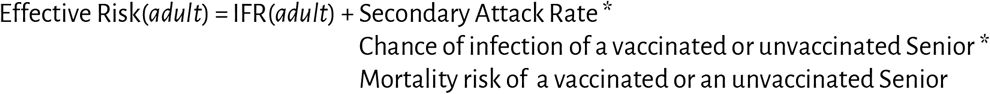

This definition is conditional to the assumption that there is a person with a primary infection in the household, similar to the definition of secondary attack rate conditional to a primary infection. We used the infection fatality rate data (**Supplementary Table 2**) from Levin *et al*. 2020 to calculate the effective risk and to estimate the effective age (**Table 1**). **Table 1** utilizes the home contact structure for India with an approximate 25 year intergenerational gap, and the effectiveness of the COVISHIELD (*Serum Institute of India – AstraZeneca Plc*) vaccines that are mostly used in India. It is evident that the “effective age” of an Adult in a multi-generational home is 15 years more if living with an unvaccinated parent (Senior) or 40 years more if living with an unvaccinated grandparent (Senior). This personal risk quantification clarifies the our estimates of the advantages of preferential vaccination of Adults from multigenerational homes as a public health strategy.

**Table 1.** Reduction of deaths among Seniors. The two different vaccination strategies employed have varying impacts on the transmission of infections from adults to Seniors, and the consequent deaths among Seniors. The results from the model are shown under the different pandemic scenarios. Also, while we assume the age-dependent mortality rate obtained from the meta-analysis of Levin *et al*. 2020. The lives saved are also considerably high even if the mortality rates may be 20% lesser in the second wave in India, due to better therapeutic management or other reasons.

| Pattern of daily new infections for 180 days | Deaths among Seniors due to transmission from adults.<br>Vaccinating 2 million/day |  | Deaths among Seniors due to transmission from adults<br>Vaccinating 3 million/day |  |
| --- | --- | --- | --- | --- |
|  | Uniform vaccination | Targeted vaccination of Adults (20-60) | Uniform vaccination | Targeted vaccination of Adults (20-60) |
| Scenario 1 | 189,652 | 85,513 | 187,901 | 54,871 |
| Scenario 2 | 183,443 | 109,663 | 182,510 | 75,077 |
| Scenario 3 | 343,988 | 119,905 | 333,952 | 76,732 |
| Scenario 4 | 363,405 | 102,709 | 350,929 | 60,460 |

**Table 2.**
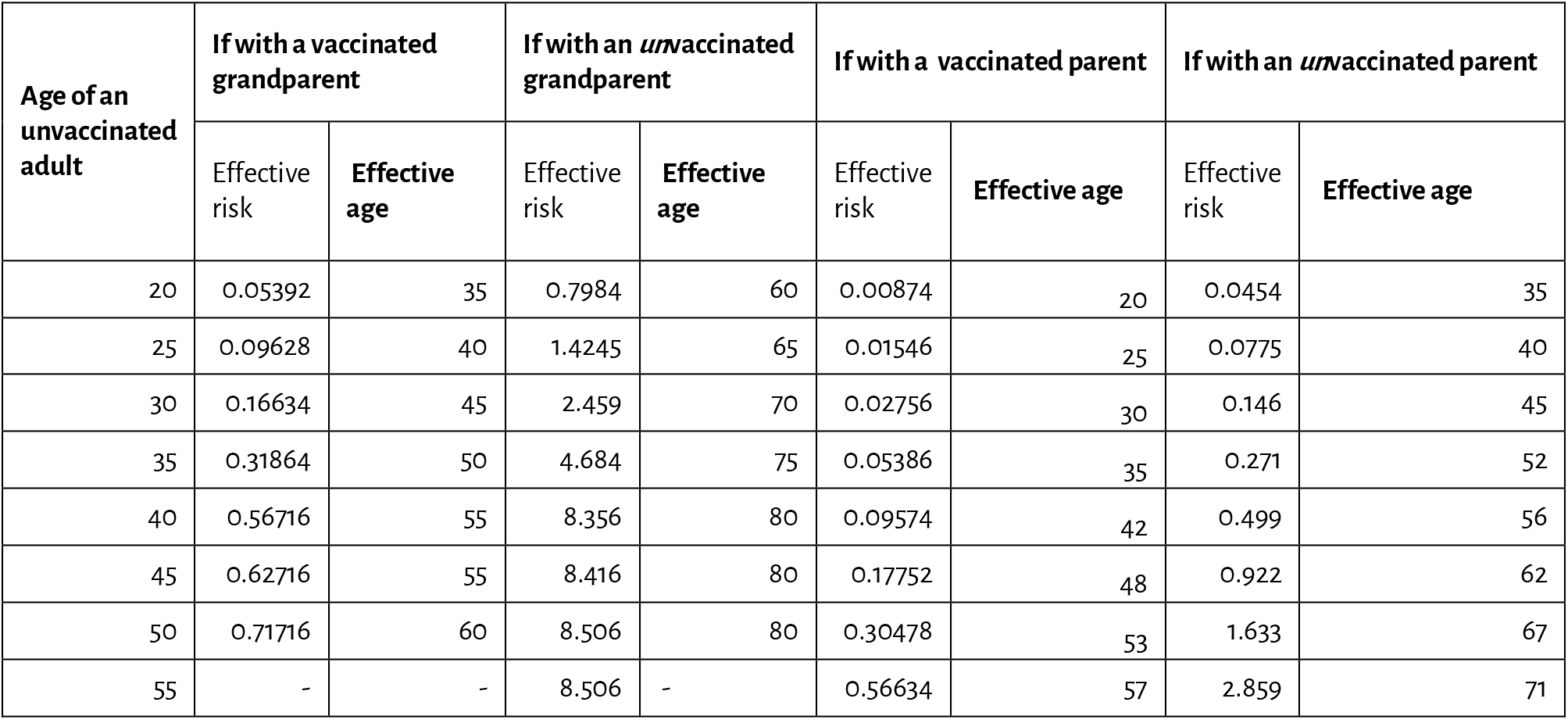
Effective Age. The “effective risk” for each adult in the age 20-55 living in a multigenerational home, is defined based on the COVISHIELD (AstraZeneca) effectiveness and a 25 year intergenerational gap. The age-dependent infection fatality rate shown in **Supplementary Table 2** was obtained from Levin *et al*. 2020. The “effective age” was obtained by a reverse-lookup of the risk from the **Supplementary Table 2**. Considering the goal of reducing COVID-19 related deaths, an adult living with an unvaccinated parent or grandparent is effectively older by 15 years or 40 years respectively.

### Using *Effective risk* for public health strategy when individual risk is low

Vaccinating the family members of the vulnerable is a well-known strategy to break the transmission chain leading up to the vulnerable. This is especially recommended if the vaccine is not safe for the vulnerable or if they are not likely to develop antibodies due to a compromised immune system. Multigenerational homes have been believed to be the major factor behind the high COVID-19 casualties in many countries, Italy for example. Whereas in the vaccination strategies of USA [NASEM, 2020], the considerations of the multigenerational homes have been important mainly to identify the vulnerable ones outside the long-term care facilities. The Swiss task force explicitly prioritized “close contacts”, household members or relatives providing care to people at risk, in priority group 3. However, the rationale, the age groups which qualify and comparison between individuals of middle-age versus those who are younger but living with older members could not be gathered easily from the public domain. Further, to the best of our knowledge, public health strategies weighing the epidemiological scenarios against the vaccine availability [Bubar *et al*. 2021], have not evaluated the societal impact of vaccinating the family members of the vulnerable, considering the *local* realities of the family structure, secondary attack rates, and the effectiveness of the available vaccines. We devise the strategy based on the “effective risk” and quantify its effectiveness using our model-based analysis. This “effective risk” or “effective age” among people of low individual risk may be extended to different demographics by combining their familial, social, or work-related interactions.

### Possible immediate strategy for prioritization in India

The vaccination drive in India is very active, and ranks very high globally in absolute numbers (132 million doses administered as of 12 April 2021). Although 41 million Seniors and 91 million Adults have been vaccinated, there are still large gaps in coverage (**Supplementary Table 3**). Delhi has a vaccination coverage of 53% among the Seniors. The coverage among Seniors varied widely across the states with a mean and standard deviation of 42% and 21%, respectively. We ask if vaccinating the vulnerable, and those next to them, is easily achievable with the available resources. The number of vaccines required to completely vaccinate the Seniors is around 77 million more. Similarly, from our estimates based on the family structure model we described, the number of adults from multigenerational homes that need to be vaccinated is 144 million more. Paradoxically, the two major factors that pose a significant hurdle to effective vaccination programs are the shortage of vaccines, and a hesitancy among some of the eligible ones. Interestingly, one of the largest states Rajasthan has achieved a vaccination coverage of 78% among the Seniors, setting a precedence for allowing other states to follow a similar model. Thus, with the stock of vaccinations India has, it is feasible to vaccinate almost all of these high risk and high ‘effective risk’ populations within 2 to 3 months.

### Limitations and Future work

The calculations work were based on the epidemiological and vaccination scenarios that looked plausible. However, this situation may change dynamically with access to newer sources of vaccines. The work assumed a 25 year intergenerational gap, that we inferred from the banded contact network of Prem et al. Other scenarios such as fully-Senior households were not considered, believing that the model nevertheless captures a majority of the family structure. However, we also believe that by including the contact with the domestic-help in some of the cases, the risk exposure to the Seniors through Adults may be equivalent to having younger family members at home. A study based on more granular family structured data, if available, will be interesting to recalibrate the effectiveness of the strategy we propose.

Needless to say, there is a scope for examining several other strategies in greater detail, such as – by recognizing that the pandemic spread is not uniform across the country, and the case load migrates [Ansumali *et al*. 2020], it is also possible to preferentially vaccinate these populations with high risk and ‘high effective risk’ in districts or states that are at the highest risk of COVID-19 infections; similarly, the two-pronged strategy of keeping the infections under control with lockdowns in the selective hotspots and vaccinating those regions rapidly.

## Conclusions

While successfully conducting large-scale vaccination drives, India faces the challenge of vaccinating a large young population. The “effective age” of an unvaccinated adult living with a grandparent we define by combining their risks, may be even 40 years more, thus underscoring the need to vaccinate those living with the vulnerable. By placing this risk through the Adults in the family, in the context of current vaccination rates, it is possible to prioritize and administer the vaccinations to the Seniors, and Adults who are in multigenerational homes, there by significantly reducing the chances of deaths in epidemiological scenarios where the second wave may last for an additional 3 to 6 months.

## Data Availability

The work is based on publicly available data. The scripts will be made available on request.

## Acknowledgements

SA and MKP thank Dr. Aloke Kumar for helpful discussions.

## Author contributions

SA and MKP conceptualized the project; BS and MKP performed calculations; SA and MKP wrote the article.

## Scripts

The scripts used in this analysis are available at https://github.com/meherpr/vaccinating-young

## Supplementary Information

**Supplementary Table 1.** The 95% confidence interval for the ratio of the 2^nd^ peak to the 1^st^ peak is this 4.3522 ±1.954. Similarly, the active case ratio between the second and the first waves is 3.8375 ±1.331 excluding the outlier France (and 7.0867 ±6.12 if include France). COVID-19 incidence data was obtained from *https://worldometers.info*

| Country | Peak daily infections |  |  | Active infections at peak |  |  | 2<sup>nd</sup> Peak width |  |
| --- | --- | --- | --- | --- | --- | --- | --- | --- |
|  | First wave | Second wave | Second wave/first wave | First wave | Second wave | Second wave/first wave | Width at half peak height (days) | Degree of restriction |
| USA | 35278 | 76,268 | 2.16 | 1127293 | 2444516 | 2.16 | 77 | Moderate |
| Brazil | 46263 | 77129 | 1.66 | 818533 | 1293839 | 1.58 | >103 (ongoing) | Low |
| France | 4852 | 56377 | 11.62 | 59336 | 1962739 | 33.08 | 28 | Strict |
| Russia | 10982 | 28616 | 2.60 | 245580 | 563754 | 2.29 | 122 | Low |
| UK | 4513 | 25283 | 5.60 | 193327 | 1013141 | 5.24 | 38 | Strict |
| Turkey | 12400 | 31787 | 2.56 | 86675 | 224245 | 2.58 | 37 | Strict |
| Italy | 6,554 | 41,195 | 6.28 | 108077 | 803591 | 7.43 | 44 | Strict |
| Spain | 8641 | 20169 | 2.33 | 138061 | 542985 | 3.93 | 47 | Strict |
| Germany | 5837 | 25441 | 4.36 | 72865 | 400245 | 5.49 | 91 | Moderate |

**Supplementary Table 2.** Age-dependent infection fatality risk obtained from Levin *et al*. 2020 was used in our analysis. This data was also used for a “reverse-lookup” of effective age when the effective risk was computed.

| Age | 20 | 25 | 30 | 35 | 40 | 45 | 50 | 55 | 60 | 65 | 70 | 75 | 80 | 85 | 90 | 95 | 100 |
| --- | --- | --- | --- | --- | --- | --- | --- | --- | --- | --- | --- | --- | --- | --- | --- | --- | --- |
| Mortality risk | 0.0064 | 0.0115 | 0.02 | 0.04 | 0.07 | 0.13 | 0.22 | 0.42 | 0.77 | 1.43 | 2.64 | 4.71 | 8.13 | 15.48 | 27.62 | 27.62 | 27.62 |

**Supplementary Table 3.** Vaccine coverage and needs. State wide vaccine coverage and needs among the Seniors (over 60 years) and the adults (20-60) estimated to be in multigenerational homes with a direct contact with the Seniors. Vaccination statistics are obtained from the Indian Government portal https://dashboard.cowin.gov.in/

| State | Seniors (age 60+) |  | Estimated adults (ages 20-60) in Family |  |
| --- | --- | --- | --- | --- |
|  | Vaccinated | Still needing vaccines | Vaccinated | Still needing vaccine |
| Andaman and Nicobar | 10,818 | 17,784 | 3,382 | 34,659 |
| Andhra Pradesh | 1,331,327 | 4,157,514 | 490,817 | 7,300,159 |
| Arunachal Pradesh | 22,490 | 49,104 | 10,831 | 84,389 |
| Assam | 427,893 | 1,910,469 | 143,789 | 2,966,232 |
| Bihar | 2,226,271 | 6,444,267 | 458,751 | 11,073,065 |
| Chandigarh | 41,381 | 34,082 | 8,638 | 91,728 |
| Chattisgarh | 1,438,098 | 816,300 | 520,177 | 2,478,172 |
| Dadra and Nagar Haweli | 6,531 | 9,098 | 2,497 | 18,290 |
| Daman and Diu | 10,311 | 2,470 | 2,094 | 14,905 |
| Delhi | 695,327 | 595,549 | 181,430 | 1,535,435 |
| Goa | 75,917 | 108,015 | 18,413 | 226,217 |
| Gujarat | 3,525,625 | 1,859,254 | 929,160 | 6,232,729 |
| Haryana | 1,154,937 | 1,313,038 | 240,115 | 3,042,292 |
| Himachal Pradesh | 453,561 | 337,324 | 112,291 | 939,586 |
| Jammu and Kashmir | 444,115 | 593,873 | 140,655 | 1,198,453 |
| Jharkhand | 967,958 | 1,683,305 | 225,656 | 3,300,524 |
| Karnataka | 2,725,660 | 3,789,251 | 684,709 | 7,980,123 |
| Kerala | 2,565,167 | 2,152,400 | 550,710 | 5,723,654 |
| Ladakh | 23,859 | 6,141 | 8,349 | 33,067 |
| Lakshadweep | 3,358 | 2,571 | 1,410 | 6,476 |
| Madhya Pradesh | 2,433,448 | 3,994,033 | 624,500 | 7,924,050 |
| Maharashtra | 4,203,234 | 8,292,068 | 1,253,221 | 15,365,531 |
| Manipur | 3,813 | 221,210 | 15,477 | 283,804 |
| Meghalaya | 26,220 | 130,045 | 11,146 | 196,686 |
| Mizoram | 25,377 | 51,830 | 8,061 | 94,624 |
| Nagaland | 19,121 | 96,446 | 8,867 | 144,837 |
| Odisha | 1,894,449 | 2,588,055 | 449,739 | 5,511,991 |
| Puducherry | 32,121 | 103,370 | 14,630 | 165,573 |
| Punjab | 696,442 | 2,527,602 | 261,600 | 4,026,379 |
| Rajasthan | 4,497,085 | 1,254,071 | 873,591 | 6,775,446 |
| Sikkim | 38,018 | 7,828 | 10,643 | 50,332 |
| Tamil Nadu | 1,237,026 | 7,211,452 | 510,430 | 10,726,046 |
| Telangana | 739,704 | 1,653,896 | 277,492 | 4,795,500 |
| Tripura | 247,641 | 78,096 | 92,798 | 340,432 |
| Uttar Pradesh | 3,380,058 | 13,989,835 | 995,726 | 22,106,232 |
| Uttarakhand | 539,872 | 473,538 | 147,081 | 1,200,754 |
| West Bengal | 3,185,711 | 5,524,469 | 810,169 | 10,774,370 |

**Supplementary Scheme 1.**
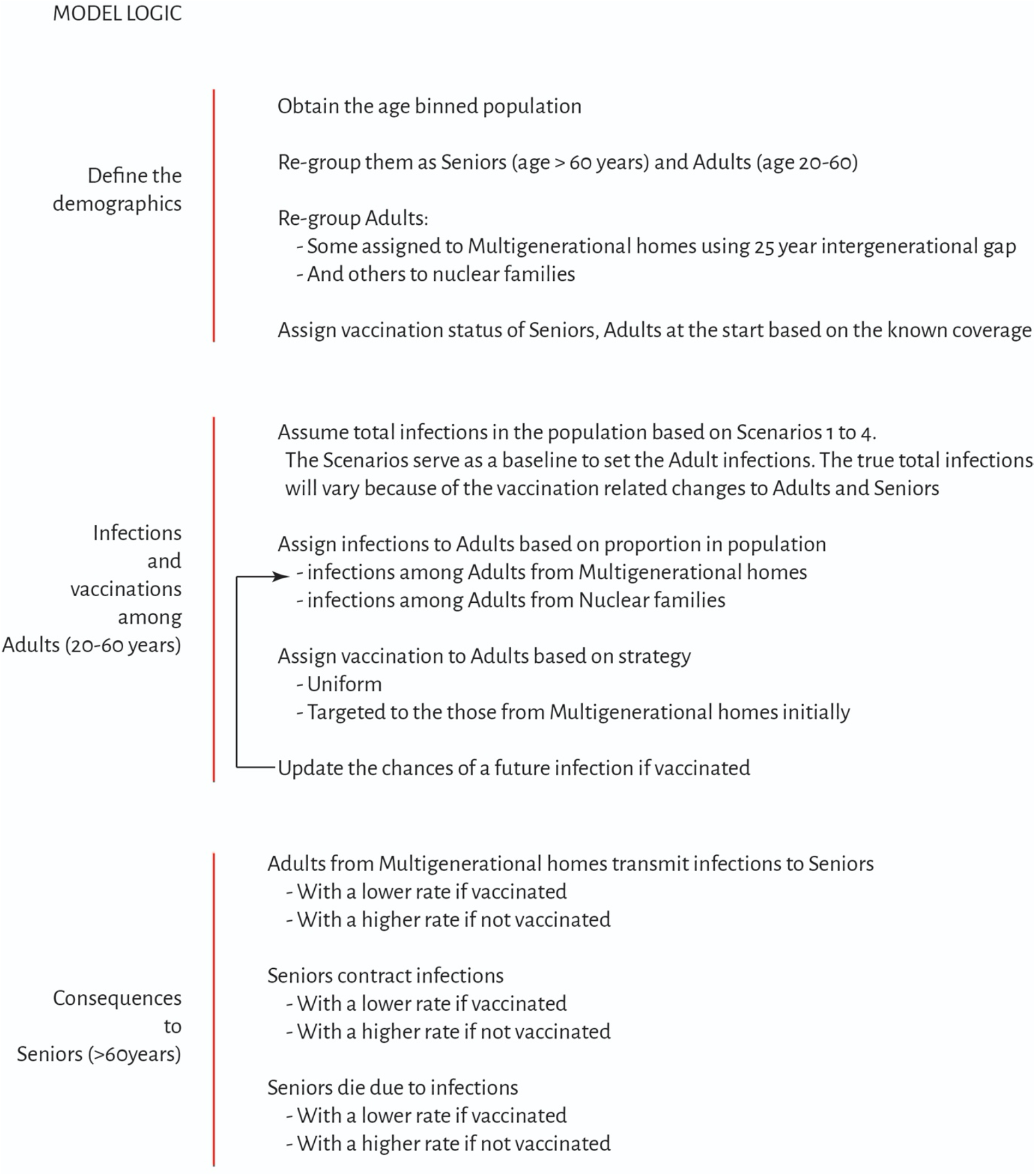
Model logic. The scheme illustrates how the calculations were performed in three stages – 1. Demographics are defined 2. Infection and vaccination status for the adults belonging to multigenerational families or otherwise are assigned. The complete code for performing the calculations is available at – https://github.com/meherpr/vaccinating-young

